# Integrating mobility, travel survey, and malaria case data to understand drivers of malaria importation to Zanzibar, 2022-2023

**DOI:** 10.1101/2025.06.04.25327997

**Authors:** Julia G. Muller, Kieran Dewalt, Varun Goel, Mohamed Ali, Wahida Hassan, Bakari Mohamed, Barbara Bella Choloi, Dominick Msolo, Abdallah Zacharia, Mwanaidi Nwange, Rokhaya Kane, Donal Bisanzio, Michael Emch, Anders Björkman, Richard Reithinger, Shija J. Shija, Billy E. Ngasala, Jonathan J. Juliano, Jessica T. Lin

## Abstract

**Background:** Zanzibar has achieved historic reductions in malaria incidence, but high connectivity to mainland Tanzania and imported cases remain a challenge to “last mile” malaria elimination.

**Methods:** To understand factors driving malaria importation, we collected travel histories and demographics of malaria cases presenting to 94 health facilities across Zanzibar’s main island, Unguja, from 2022–2023. We also analyzed population mobility data—self-reported travel at the outbound Dar es Salaam ferry terminal and Meta colocation and movement distribution—to examine movement patterns between Unguja and mainland Tanzania. We integrated these with Climate Hazards Group InfraRed Precipitation with Station (CHIRPS) rainfall data to explore the seasonality of human movement and travel-associated malaria risk.

**Results:** Among 1,172 malaria cases reporting recent travel to mainland Tanzania, travel to Tanga (20%), Dar es Salaam (20%), and Morogoro (15%) were most common. Nearly half of travelers had spent over 14 nights away from home; the majority were visiting family. While mainlanders made up two-thirds of ferry travelers, 28% of travel-associated malaria cases in Unguja reported primary residence on the mainland. Zanzibari residents who reported travel to mainland regions with high/moderate malaria risk during the dry season composed the highest proportion of travel-related malaria cases. Long-distance travel off Unguja decreased during the rainy seasons, and imported cases correlated with rainfall at the mainland source, rather than in Zanzibar. With different biases, colocation and ferry data approximated the proportional makeup of at-risk travel relatively well.

**Conclusions:** Movement flows and seasonal rainfall patterns drive imported malaria in predictable ways that can be harnessed to target high-risk travelers for intervention.

## BACKGROUND

Zanzibar, a semi-autonomous archipelago within the Republic of Tanzania, achieved historic reductions in malaria incidence in the decade following 2002 through extensive interventions [1,2]. The Zanzibar Malaria Elimination Program (ZAMEP) sustained these reductions through ongoing case-based surveillance and active follow-up of every malaria case diagnosed across the islands [3,4]. However, malaria resurgences in 2020 and 2023 highlighted the fragility of these hard-won gains and the challenges the region faces in pursuing malaria elimination [1,2,5,6]. Key among these challenges is importation of malaria from higher transmission areas, especially mainland Tanzania [1,7–10]. Zanzibar lies 36.5km from the coast of Tanzania, with ferries transporting more than 18,000 passengers daily between Dar es Salaam, the main coastal hub, and Unguja, the largest and most populous island in Zanzibar [8,11]. Thus, like many other areas of sub-Saharan Africa trying to achieve “last mile” malaria elimination, the region remains vulnerable to its re-introduction through human migration. Understanding the contours of human migration, their seasonal variation, and their relationship to malaria cases reported in Zanzibar is crucial for informing targeted interventions [10].

Human mobility data have proven valuable for mapping the distribution and movement patterns of populations vulnerable to malaria [12–14]. Pioneering work used mobile phone and ferry traffic data to measure malaria-relevant human movement flows in Zanzibar, highlighting the importance of returning Zanzibari travelers to malaria importation [8,9]. Zanzibar’s mobile phone market is now highly fragmented, and privacy concerns have hindered access to cellular data for mobility analysis [15]. As an alternative, we used colocation and movement distribution data provided by Meta (previously Facebook) Data for Good (DFG). These data have been widely used, especially during the COVID-19 pandemic [16]. We also obtained our own mobility data through surveys administered to ferry travelers at the outgoing Unguja-bound ferry terminal in Dar es Salaam.

We integrated this mobility data, as well as publicly available rainfall data, with comprehensive Zanzibar case data obtained from health facilities in 2022-2023 as part of our larger Zanzibar Imported Malaria (ZIM) project [17]. Overall, this study aims to provide an updated understanding of the role of human mobility and seasonality in shaping travel-associated malaria in Zanzibar. We first identify factors associated with imported malaria in Unguja. We then examine how rainfall patterns in Unguja and in mainland Tanzania affect human mobility and malaria case incidence in Unguja. Finally, the Meta colocation data and ferry survey data are used to describe movement patterns between mainland Tanzania and Zanzibar, focusing on differences between mainland and Zanzibari travelers, and comparing the ability of these different data sources to estimate the proportion of malaria importation arising from different mainland regions.

## METHODS

Within the Zanzibar archipelago, Unguja (6°08’26”S, 39°20’12”E) has historically borne a higher malaria burden, partly due to being the main entry point for travelers. Unguja’s climate is characterized by two primary rainy seasons—*masika* (March-May) and *vuli* (October-December)—each followed by a peak in malaria transmission.

Prospective surveys were carried out among malaria cases presenting to health centers in Unguja and ferry travelers bound for Unguja with approval from the institutional review boards (IRB) in Zanzibar (ZAHREC/01/EXT/July/2023/03), at MUHAS (MUHAS-REC-02-2024-1168), and at UNC (21-1966). Written informed consent, parental consent and/or assent were provided by all participants.

### Health facility surveys

Clinic surveys were administered to malaria cases diagnosed by rapid diagnostic test (RDT) at clinics in Unguja from May 2022 to December 2023. A total of 94 health facilities on Unguja were selected using a stratified random sampling approach to draw evenly from urban and rural areas (Supplementary Table 1). Data were collected through standardized Case Report Forms (CRFs), administered by the health center staff, with training and oversight by ZAMEP malaria surveillance officers. CRFs collected information on demographic factors like age, sex, occupation, and home district, as well as travel history, including (up to two) primary travel destinations, duration of travel, and reason for travel. Survey responses were scanned into an electronic database using ABBYY Flexcapture software (ABBYY Corporation, TX). Data were manually checked and corrected as needed by research staff.

### Ferry surveys

Ferry surveys were conducted among ticketed outbound travelers at the Dar es Salaam ferry terminal—including Zanzibaris returning home and mainlanders traveling to Zanzibar—between August and December 2024. Data in this report were drawn from all those screened for survey participation, in which travelers were asked to report their home region/district, whether they were leaving or returning home, travel destination (region/district), number of nights spent away, and number of people traveling with them.

Responses were collected manually and entered into Microsoft Excel. Reported regions and districts were matched with those listed in the Database of Global Administrative Boundaries (GADM) [18].

### Meta colocation data

Colocation data from mainland Tanzania and Unguja spanning December 2022-September 2024 were downloaded from the Meta DFG portal. Meta’s methodology for collecting this data has been published previously [16]. Data include weekly estimates of the probability that a randomly selected individual from one district and a randomly selected individual from another district were in the same location—specifically, the same level 16 Bing tile. These estimates are provided for each pair of districts and represent the degree of interaction between them. Further details on our calculations and approach to data cleaning are provided in the Supplement. For each Zanzibar-mainland district pair, we calculated the average over time of the observed colocation and observed co-observation, then calculated their quotient as the district-district colocation rate.

This was multiplied by both districts’ population in Tanzania’s 2022 census [19] to give weighted colocation. The district-district weighted colocation was summed over all districts in Unguja and districts within a mainland region to obtain an estimate for the total interaction between all of Unguja and each mainland region.

### Meta movement distribution data

Movement distribution data provide a daily proportion of Facebook users with location services enabled who travel a maximum Euclidean distance from their home district within 4 categories: 0 km, 1-9 km, 10-100 km, and >100 km. To aggregate over a group of districts, the proportions were multiplied by the district population to estimate the number of people in the category, followed by summation of the estimated counts across districts in each group.

### Statistical Analysis

From the health center surveys, we determined the top destinations for those who reported travel to the mainland in the previous 28 days. We compared demographic characteristics of cases who did and did not report recent travel, and factors associated with short- and long-term travel (defined as 14 or fewer vs. greater than 14 nights away), using chi-square or Fisher’s exact tests, as appropriate.

Using Meta movement distribution data, we compared the proportion of travelers traveling long (>100 km) and medium (10-100 km) distances on rainy vs. dry days (cumulative rainfall over the previous 30 days of greater or less than 100 mm, respectively), using Mann-Whitney U-testing. Data on regional rainfall were obtained from Climate Hazards Group InfraRed Precipitation with Station (CHIRPS) data [20] through Google Earth Engine [21]. We also modeled the relationship between weekly malaria case counts for travelers from each travel destination region and rainfall at that region using negative binomial regression. Cumulative rainfall from 2-6 weeks prior to clinic visit date was calculated to account for vector ecology, the incubation period of malaria, and median travel time [22].

Finally, we compared proportions of travelers arriving from common travel destinations based on data derived from Meta colocation data, ferry screening, and Zanzibar malaria clinic cases. Results were visualized in proportion plots which compared relative proportions of travelers captured by each data source. Region- and district-level malaria risk strata were obtained from Tanzania’s 2021-2025 National Malaria Strategic Plan [6] (Supplementary Figure 1). For regional analyses, Kagera, Mwanza, Geita, Shinyanga, Simiyu, and Mara were grouped as the “Lake Zone” [23]. For all analyses, significance was tested at an ? level of 0.05, with no adjustment made for multiple testing. Analyses were performed using R (4.4.2), Python (3.10.14), and Tableau Public (2024.3.0).

## RESULTS

### Factors associated with travel-related malaria risk

Among 3,875 individuals who presented to clinics in Unguja with malaria from May 2022 to December 2023, 1,172 (30%) reported travel to mainland Tanzania within the past 28 days and were presumptively categorized as imported or travel-associated malaria cases. The majority of imported cases were reported from central Unguja, including the urban districts of Mjini and Magharibi. Travelers most often visited Tanga (n=238, 20%), Dar es Salaam (n=231, 20%), Morogoro (n=170, 15%), and Pwani (n=137, 12%) regions; 144 cases (12%) reported traveling to the Lake Zone (Figure 1).

**Figure 1.**
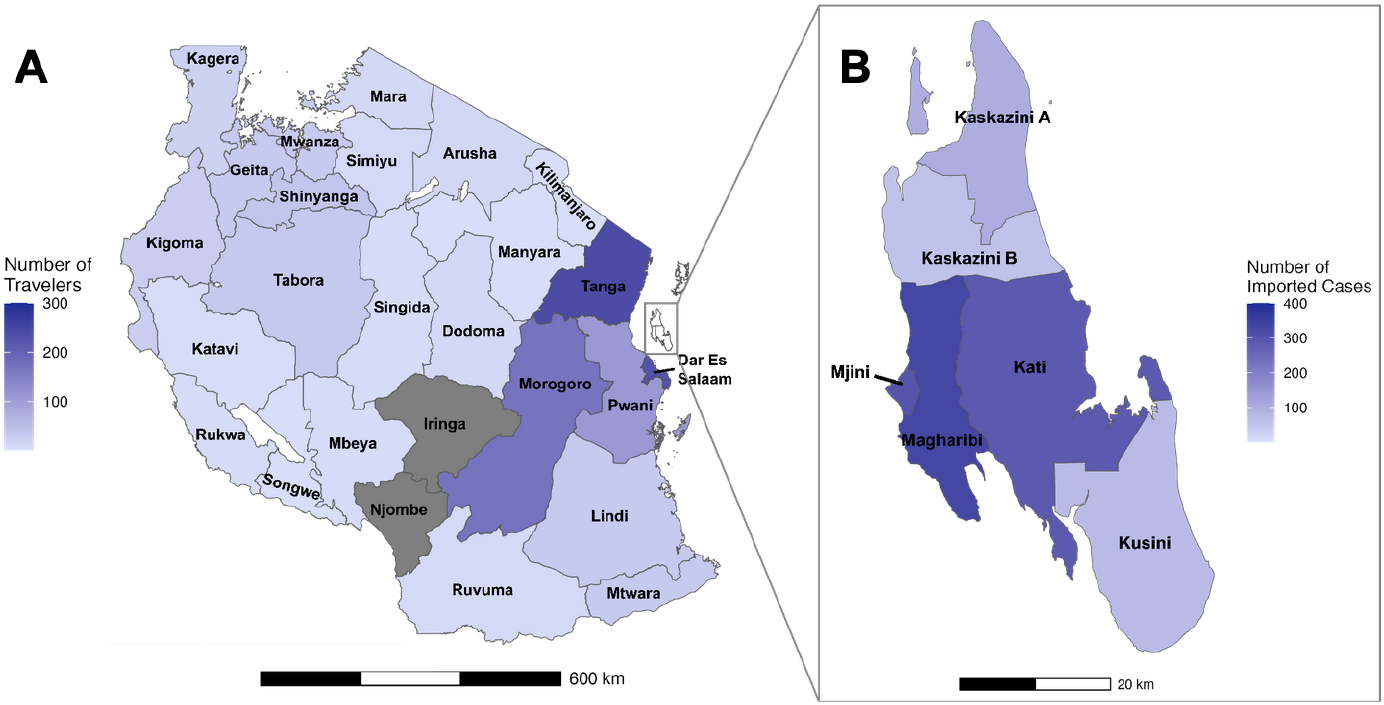
(A) Self-reported travel destinations for confirmed malaria cases on Zanzibar who reported recent travel to the mainland and (B) home districts of travel-associated malaria cases on Zanzibar, May 2022 to December 2023 (n=1,172). Darker blue indicates more travelers, while lighter blue represents fewer travelers. Mainland regions shown in dark gray indicate no recorded travel from Zanzibar.

Compared to non-travel-related malaria cases, a higher proportion of imported cases were in children under five years of age (11.1% in travelers vs. 4.0% in non-travelers, *p*<0.01) (Table 1). In terms of occupational status, a higher proportion of travel-related cases was observed in farmers (22.6% vs. 10.4%, *p*<0.01) and housewives (14.8% vs. 10.0%, *p*<0.05). In contrast, a lower proportion of travel-related cases was observed in night watchmen/security and students. However, night watchmen were more prevalent among cases reporting travel within Zanzibar (Supplementary Table 2).

**Table 1.** Select demographic characteristics of Zanzibari confirmed malaria cases (overall and by self-reported recent travel to the mainland), May 2022 to December 2023. The table includes self-reported sex, age, and occupation of Zanzibari cases. Chi-square comparisons are included to assess the differences between travelers and non-travelers across these characteristics. Significant values (p<0.05) are highlighted in bold.

|  | Total<br>(N=3875) | Non-travelers<br>(N=2703) | Travelers<br>(N=1172) | P-value |
| --- | --- | --- | --- | --- |
| **Sex** |  |  |  |  |
| Female | 1265 (32.6%) | 776 (28.7%) | 489 (41.7%) | **<0.001** |
| Male | 2604 (67.2%) | 1925 (71.2%) | 679 (57.9%) |  |
| **Age** |  |  |  |  |
| Under 5 | 238 (6.1%) | 108 (4.0%) | 130 (11.1%) | **<0.001** |
| 5 to 14 | 438 (11.3%) | 287 (10.6%) | 151 (12.9%) |  |
| 15 to 24 | 1345 (34.7%) | 968 (35.8%) | 377 (32.2%) |  |
| 25 to 39 | 1227 (31.7%) | 879 (32.5%) | 348 (29.7%) |  |
| 40 and older | 627 (16.2%) | 461 (17.1%) | 166 (14.2%) |  |
| **Occupation\*** |  |  |  |  |
| Trader/business | 762 (25.4%) | 543 (25.2%) | 219 (25.8%) | 0.60 |
| Farming | 416 (13.9%) | 224 (10.4%) | 192 (22.6%) | **<0.001** |
| Housewife | 341 (11.4%) | 215 (10.0%) | 126 (14.8%) | **0.02** |
| Watchman/security | 233 (7.8%) | 213 (9.9%) | 20 (2.4%) | **<0.001** |
| Student | 211 (7.0%) | 176 (8.2%) | 35 (4.1%) | **<0.001** |
| Fishing | 182 (6.1%) | 138 (6.4%) | 44 (5.2%) | 0.19 |
| Public servant/NGO | 131 (4.4%) | 97 (4.5%) | 34 (4.0%) | 0.55 |
| Tourism | 96 (3.2%) | 65 (3.0%) | 31 (3.7%) | 0.91 |
| Food service | 95 (3.2%) | 70 (3.3%) | 25 (2.9%) | 0.70 |
| Construction | 48 (1.6%) | 34 (1.6%) | 14 (1.6%) | 0.99 |
| Other | 487 (16.2%) | 378 (17.6%) | 109 (12.8%) | **<0.001** |
| Child | 830 | 520 | 310 |  |
| Missing | 43 | 30 | 13 |  |

\*Percentages are based on totals minus children and those missing occupation. Only occupations with >1% frequency are shown.

More than half of travel-associated malaria cases (52%, 520/1001) reported more than 14 nights spent away from home. In contrast, only 26% (103/389) of ferry travelers returning home to Zanzibar reported spending more than 14 nights away (Supplementary Figure 2). These long-term travelers were more likely to have visited mainland regions considered to be at high or moderate risk for malaria (Table 2). Visiting family was the leading reason for travel among both long- and short-term travelers.

**Table 2.**
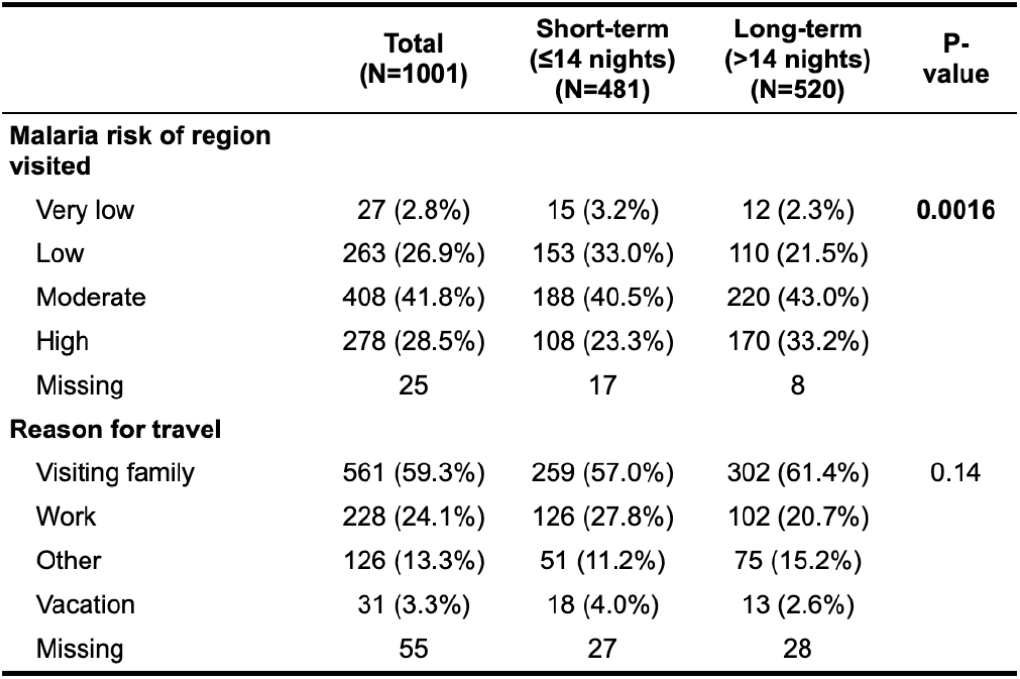
Malaria risk of region visited and self-reported reason for travel for Zanzibar confirmed malaria cases who reported recent travel to the mainland, 2022-2023. Chi-square tests were used to assess the association between travel duration and malaria prevalence. Malaria risk data is sourced from the 2021-2025 Tanzania National Malaria Strategic Plan. Travelers missing data on duration of travel are excluded from this table.

|  | Total<br>(N=1001) | Short-term<br>(≤14 nights)<br>(N=481) | Long-term<br>(>14 nights)<br>(N=520) | P-value |
| --- | --- | --- | --- | --- |
| **Malaria risk of region**<br>**visited** |  |  |  |  |
| Very low | 27 (2.8%) | 15 (3.2%) | 12 (2.3%) | **0.0016** |
| Low | 263 (26.9%) | 153 (33.0%) | 110 (21.5%) |  |
| Moderate | 408 (41.8%) | 188 (40.5%) | 220 (43.0%) |  |
| High | 278 (28.5%) | 108 (23.3%) | 170 (33.2%) |  |
| Missing | 25 | 17 | 8 |  |
| **Reason for travel** |  |  |  |  |
| Visiting family | 561 (59.3%) | 259 (57.0%) | 302 (61.4%) | 0.14 |
| Work | 228 (24.1%) | 126 (27.8%) | 102 (20.7%) |  |
| Other | 126 (13.3%) | 51 (11.2%) | 75 (15.2%) |  |
| Vacation | 31 (3.3%) | 18 (4.0%) | 13 (2.6%) |  |
| Missing | 55 | 27 | 28 |  |

A sizable proportion of Zanzibar malaria cases were found in those with strong ties to the mainland. Among all Unguja malaria cases surveyed, 15% (600/3,875) of all cases and 28% (333/1,172) of travel-associated cases reported their primary residence to be on the mainland. Almost two-thirds (372/600, 62%) of mainland cases reported having resided less than 6 months on Zanzibar; 56% (333/600) reported visiting the mainland within the past month (Supplementary Table 3). Compared to Zanzibaris with malaria, mainland cases were more likely to report farming as their primary occupation (22% of mainlanders vs. 15% of Zanzibaris, *p*<0.01), especially among those who had resided in Zanzibar for less than 6 months (24%) or recently traveled to the mainland (28%).

### Rainfall effects on mobility and malaria in Zanzibar

Imported malaria cases appeared to peak outside of the rainy season months in Zanzibar (March-May and October-December) during our study years (Supplementary Figure 3). One reason for this may be a negative effect of rainfall on human mobility that drives travel-related malaria exposure. Based on movement distribution data from December 2022 to August 2024 totaling 107,683 daily observations, 3-7% of Zanzibari Facebook users traveled medium distances (10-100km) on a monthly basis (Figure 2A). This distance includes movement between districts within Zanzibar or to nearby coastal cities on the mainland, such as Dar es Salaam. An even smaller fraction, only 0.2-1.2% of Facebook users in Zanzibar, traveled more than 100km on a monthly basis (Figure 2C). Both medium- and long-distance travel decreased during the long rainy season (March-May), especially from urban districts, where travel generally surpassed that in rural districts. Less pronounced dips in both types of travel were also observed during the short rainy season (October-December). Overall, both long- and medium-distance travel were lower on days when the previous 30 days of accumulated rainfall surpassed 100mm (*p*<0.01 for both comparisons).

**Figure 2.**
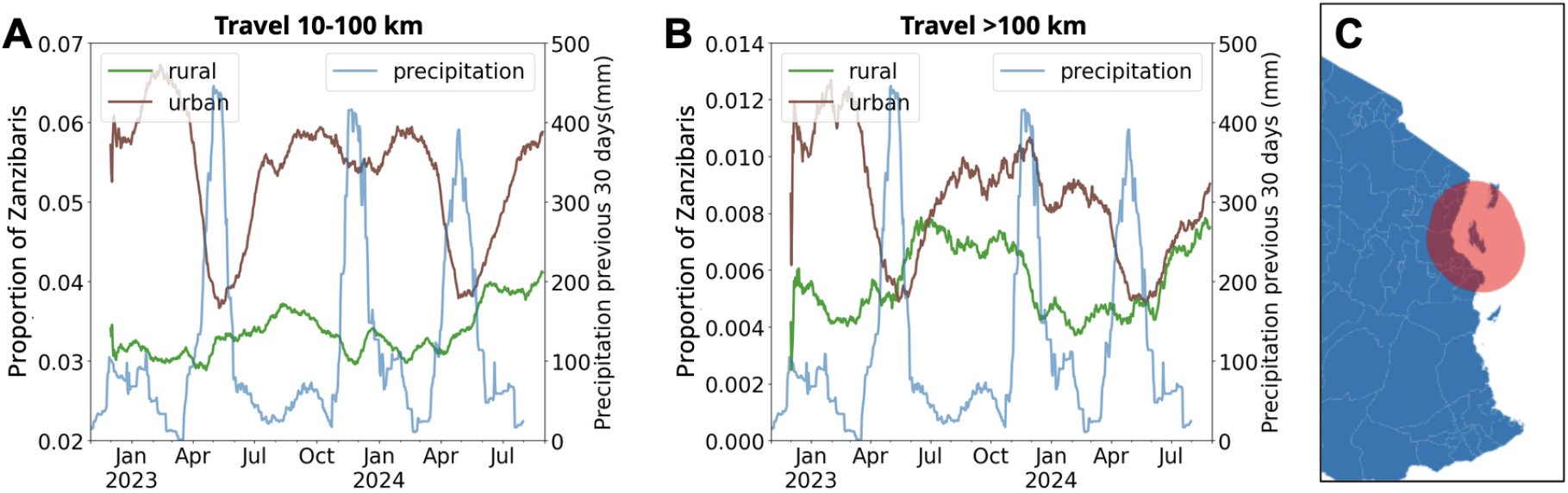
Proportion of Zanzibaris traveling (A) medium distances (10-100 km) or (B) long distances (>100 km) from their home district (classified as rural or urban), plotted alongside cumulative precipitation over the previous 30 days from 2023 to 2024. Urban districts include Mjini and Magharibi; rural districts include Kaskazini A & B, Kusini, and Kati. Left y-axes show the proportion of Zanzibaris traveling while the right y-axes show precipitation in mm. Travel estimates are based on anonymized location data from Facebook users in Unguja with location services enabled. (C) Map showing a 100-km radius around Unguja (shaded in red), encompassing common medium-distance destinations such as Dar es Salaam and coastal parts of Tanga and Pwani regions.

Another reason that imported malaria cases in Zanzibar may peak outside the rainy season is the rainfall patterns of mainland Tanzania differ from that in Zanzibar. Imported malaria case counts were associated with rainfall in the mainland regions visited, with a 6% increase in cases per inch increase in rainfall (IRR: 1.06, 95% CI: 1.04, 1.08). Meanwhile, non-travel-associated malaria cases on Zanzibar were associated with recent rainfall in Zanzibar, with a 10% increase in cases per inch increase in rainfall (IRR: 1.10, 95% CI: 1.05, 1.15) (Figure 3).

**Figure 3.**
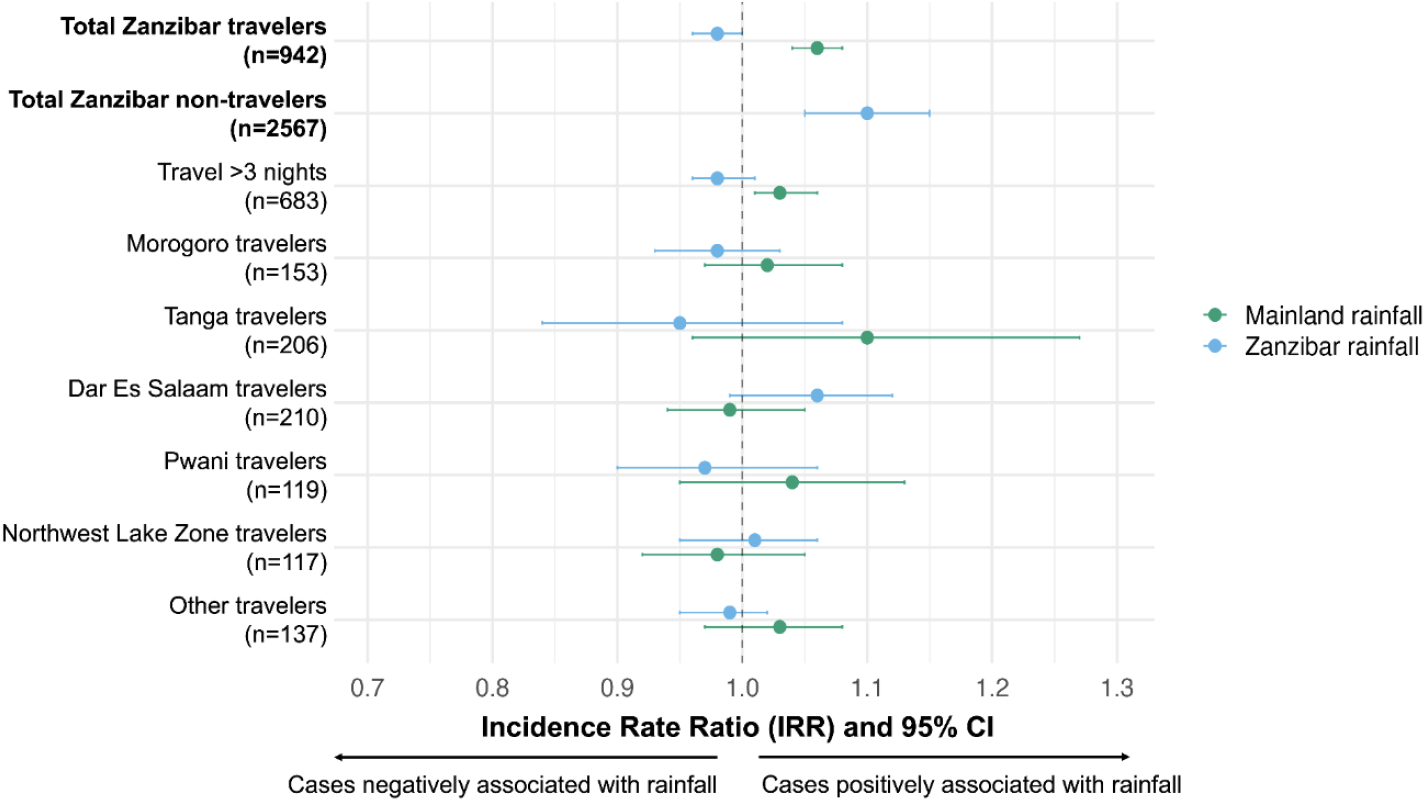
Association between malaria cases in Zanzibar and regional rainfall. The figure presents the results of negative binomial regression models evaluating the association between malaria case counts in Zanzibar and cumulative regional rainfall (Zanzibar and mainland Tanzania) for cases occurring from October 2022 onward. A 2-week lag prior to clinic visit was applied for each case, and rainfall was summed over the preceding 4 weeks. Malaria cases were grouped by the travel region visited, and the models assessed ther elationship between case counts and rainfall in the corresponding region. Each point depicts the incidence rate ratio (IRR) with its 95% confidence interval (Cl). Associations with mainland rainfall are shown in green, and those with Zanzibar rainfall are in blue. Results are shown for all Zanzibar travelers and non-travelers, as well as for key subgroups: travelers spending more than 3 nights at their destination and those visiting common destinations.

### Mainland-Zanzibar travel patterns

Meta DFG colocation data and ferry traveler survey data differed in their depiction of human movement between mainland Tanzania and Zanzibar (Figure 4). Colocation data, which depicts population mixing among mobile phone owners with Facebook location services turned on, measured 18,651 weekly colocation estimates between a district in Unguja and another district in Tanzania from December 2022 to September 2024. Regions along the eastern coast and northeast border of Tanzania, including Dar es Salaam, Tanga, Kilimanjaro, Pwani, and Arusha, were the top mainland regions exhibiting interactions with Zanzibaris (Figure 4A). In terms of interactions with individual districts within Unguja, even though the main ferry terminal on the mainland is in Dar es Salaam, the top-ranking mainland-Unguja colocation district pairs were Pangani-Mjini, Mwanga-Kati, and Pangani-Kazkazini A (Supplementary Figure 4). Mwanga, in Kilimanjaro, is in a low malaria risk region. However, Pangani is a fishing port in Tanga where ferries and boats can directly travel to Zanzibar City (Mjini) as well as to official and unofficial ports in Kazkazini A. Colocation interactions did not differ in rainy compared to dry season months (Supplementary Figure 5).

**Figure 4.**
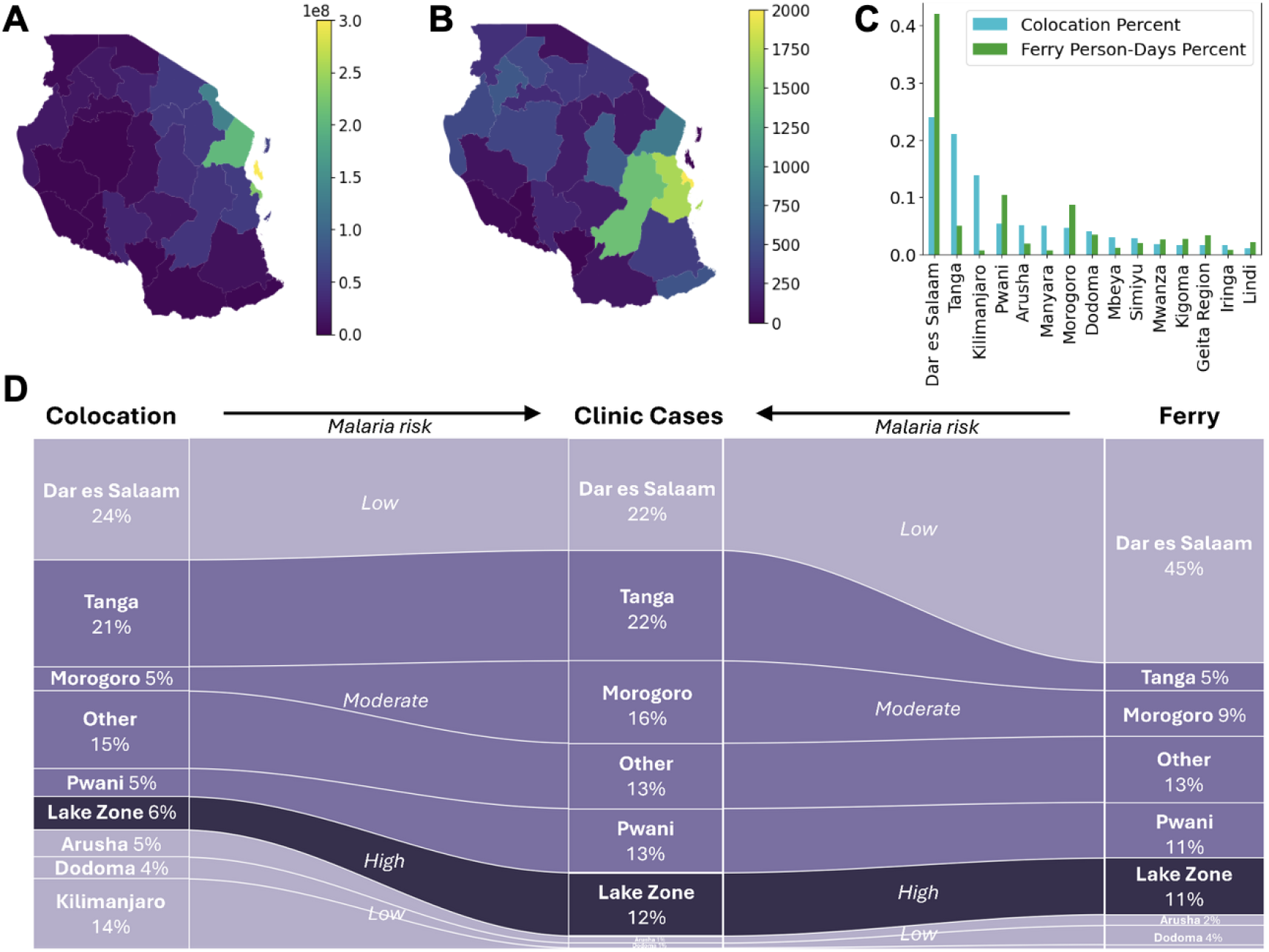
Estimation of human mobility between Unguja and mainland Tanzania. **(A)** Weighted colocation between Unguja and mainland Tanzania, aggregated between December 2022 and September 2024. Colocation represents the probability that individuals from these regions are within a small distance of one another, weighted by population. **(B)** Origins and travel destinations of ferry passengers traveling from Dar es Salaam to Unguja in August-September and November-December 2024, weighted by the length of travel (up to 28 days). **(C)** Comparison of the percent contribution of regions to travel between the mainland and Unguja, according to both the colocation data and ferry survey data. **(D)** Relative proportions of Zanzibar travelers captured through colocation data (left) and ferry surveys (right), compared to travel-associated malaria clinic cases (middle). Colors represent regional malaria risk, with darker shades indicating higher risk. Depicted risk is an average of region-level malaria risk strata (from the 2021-2025 Tanzania National Malaria Strategic Plan), weighted by population. Note that travel to Kilimanjaro is not represented for clinic cases or ferry travelers, as it accounts for less than 1% of travel volume.

Unlike colocation data, ferry survey data provides directionality of travel. Among 1,184 travelers screened at the Unguja-bound ferry terminal in Dar es Salaam between August-September and November-December 2024, two-thirds were mainlanders traveling to Zanzibar (n=795, 67%) while one-third were Zanzibaris returning home (n=389, 33%). Top mainland origins/travel destinations, weighted by duration of travel capped at 28 days, were Dar es Salaam (7189, 43%), Pwani (1752, 10%), Morogoro (1437, 8.6%), Tanga (850, 5.1%), and Dodoma (579, 3.5%), where the total person-travel days among all travelers was 16,769 (Figure 4B). A greater proportion of mainlanders vs. Zanzibaris were traveling from Morogoro, Tanga, Mtwara, and Geita, while a greater proportion of Zanzibaris were returning from Pwani and Dodoma (Supplementary Figure 6).

Overall, the colocation data highlights population mixing in coastal and northern regions, whereas ferry surveys show travel arriving to Zanzibar from coastal as well as more interior regions. (Figure 4C).

### Linking human mobility to malaria importation

Examining how well the colocation and ferry data predict the origin of malaria cases in Unguja highlights the role of malaria endemicity and modes of travel. Travel to mainland regions with the lowest malaria endemicity (Kilimanjaro, Arusha) does not translate to malaria cases while travel to regions with high malaria risk (Lake Zone) is overrepresented in the clinic cases, relative to the colocation-derived population mixing, and to a lesser degree, importation on the ferry (Figure 4D). The ferry data better reflects travel-related malaria from the coastal Pwani and neighboring Morogoro region. However, travel from Dar es Salaam is substantially overrepresented in the ferry data compared to the proportion of clinic cases reporting travel from Dar es Salaam (45% ferry vs. 22% clinic). Also, at-risk travel to Tanga appears to be underestimated by the ferry data (5% ferry vs. 22% clinic). The overall colocation data better depicts at-risk travel from Tanga; however, imported cases in the northern part of Unguja (Kazkazini A&B districts) associated with travel to Tanga are still substantially underestimated in the colocation data (Supplementary Figure 7). Compared to mainlanders on the ferry, there were fewer Zanzibaris visiting the Lake Zone and Morogoro; however, more of this travel appeared to translate into imported malaria cases from these regions (Supplementary Figure 8).

Overall, taking into account travel duration, primary residence (mainland vs. Zanzibar), seasonality of travel (wet vs. dry), and malaria risk of travel destination (moderate/high vs. low), the highest proportion of travel-related malaria cases was seen in Zanzibari travelers, both long- and short-term, who reported travel to mainland districts with high/moderate malaria risk in the dry season (Figure 5). Among surveyed ferry travelers, only 2.1% (19/899) fell into this high-risk category, despite accounting for 32% (309/977) of imported malaria cases.

**Figure 5.**
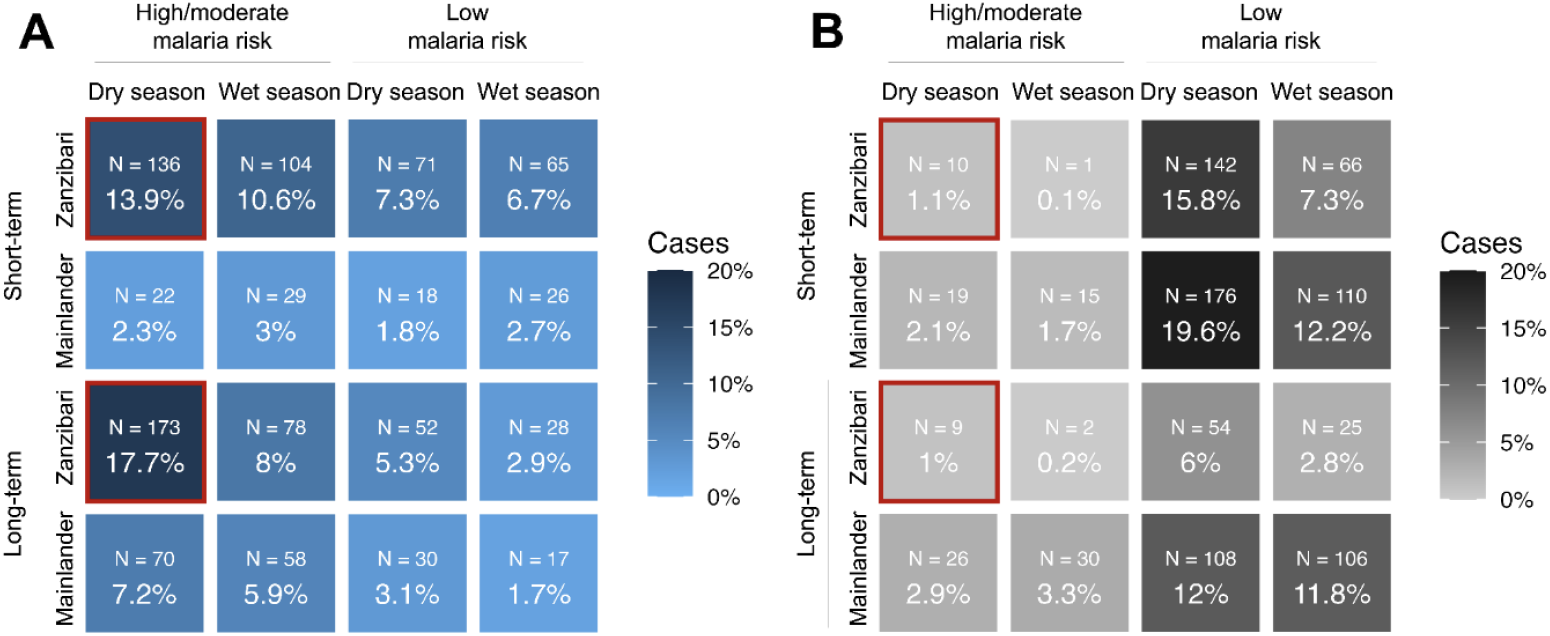
Proportions of travelers by key demographic and travel-related factors. The heat maps display the proportions of (A) Zanzibari malaria cases who reported recent travel to the mainland and (B) travelers at the outbound Dar es Salaam ferry terminal. Both heat maps are categorized by four factors: malaria transmission risk of the mainland district visited, Zanzibar season during travel (wet/dry), duration of travel (short/long; long defined as greater than 14 nights at the destination), and self-reported home residence (Mainlander vs. Zanzibari). Each cell represents the percentage of total travelers corresponding to specific combinations of these factors. Target populations of interest are marked in red.

## DISCUSSION

Human movement between Zanzibar and mainland Tanzania will likely continue to increase, keeping Zanzibar susceptible to malaria importation. This study describes imported malaria cases to Unguja in 2022-2023 and links these case data to trends in human mobility and rainfall seasonality to understand the drivers of malaria importation that can inform future intervention. Our data confirm that a few mainland regions disproportionately contribute to imported malaria to Zanzibar due to their higher malaria endemicity and connectivity to Unguja. In particular, Tanga, Pwani, Morogoro, and the Lake Zone contribute disproportionately. Importantly, human mobility data, especially those from the Meta DFG platform, approximate the composition of at-risk travel relatively well. As might be expected, traveling for longer duration, often to visit family in these regions, and bringing younger family members, poses greater risk.

Zanzibari residents make up the bulk of imported malaria cases, but young men migrating from the mainland to work and farm in Zanzibar who maintain ties and travel back to the mainland are also a key risk population.

Mobile and migrant populations present challenges to malaria control due to their diverse movement patterns and heightened exposure to malaria vectors, often coupled with limited access to interventions [24–26].

Malaria outbreaks in urban districts in Unguja in 2023 were linked to night watchmen/police, construction workers, and farmers, occupations commonly reported by mainlanders we surveyed (Supplementary Table 3) [27,28]. While research conducted 15 years ago estimated that Zanzibari residents traveling to malaria endemic regions contributed up to 15 times more imported cases than infected mainland visitors [9], this pattern may be shifting as more mainland residents travel to Zanzibar over time [11]. It is also plausible that mainlanders—who travel to regions with higher malaria risk for longer periods of time (Supplementary Figure 6)—are more likely to be semi-immune, leading them to seek care less frequently but still contribute to local transmission via chronic asymptomatic carriage [29,30].

The seasonality of at-risk travel is likely complex. Meta movement distribution data showed a negative association between local rainfall and travel, especially from urban districts where many travelers reside. However, colocation data did not show seasonal shifts in the patterns of interaction between mainland regions and Unguja. Disparate rainfall patterns at source/destination regions likely drive exposure to malaria and the timing of imported cases. These findings may help explain previously observed patterns, including increased malaria risk associated with travel during the dry season (July-September) [10], and a 12-13 week lag in cross-correlation analysis of weekly precipitation on Zanzibar and weekly reported index cases [3].

Our findings support a strategy targeting high-risk travelers for preventive interventions to reduce malaria importation [31–35]. In a 2017-18 study using reactive case detection data, travelers to regions of high malaria endemicity contributed to 77% of all travel-related infections. Our health facility data similarly highlights the outsized contribution of travel to more endemic areas. Although mainlanders outnumbered Zanzibari travelers 2.2 to 1 in our surveys on the ferry, Zanzibari travelers account for more imported malaria cases (2.6 to 1) (Supplementary Figure 7). Targeting Zanzibari travelers traveling to moderate/high-risk malaria regions in the dry season months could mean intervening on <5% of ferry travelers while potentially preventing up to one-third of imported malaria cases (Figure 5). This could be presumptive treatment or prophylaxis at ports of departure or entry [25,36].

Prior modeling work suggests that border post interventions are most effective in near-elimination settings like Zanzibar, especially when targeting major travel entry points [37]. Our data highlight Dar es Salaam as a primary travel hub and destination linking Unguja and mainland Tanzania. However, we also observed substantial travel and malaria importation from Tanga, likely arriving via vessels to official and unofficial ports in Kaskazini A [11]. Thus, digital mobility data can highlight populations underrepresented using traditional survey methods that would need to be considered in any proposed intervention [9,37].

Our study has some key limitations. First, imported cases were categorized based on travel within the previous 28 days, but residual local transmission may have led to some of these cases being acquired locally, especially if travel was limited to Dar es Salaam. Second, we lacked a control group of travelers without malaria to be able to directly assess travel factors associated with malaria infection. Instead, we leveraged ferry travel and digital mobility data to compare overall travel patterns to those of travel-associated malaria cases.

Reassuringly, Fakih et al., using reactive case detection data to compare qPCR-positive vs. negative travelers, generated findings that mirror and complement ours [10]. Third, we are unable to quantify bias introduced by use of Meta DFG data [38]. Meta does not release numbers of users; all data are provided as proportions.

Thus, we cannot speculate about the representativeness of the sample to the general Zanzibari population or attempt to correct for potential biases. However, social media uptake is highest in young adults, the major malaria risk group in Zanzibar [15]. Additionally, global migration flows estimated using Meta data have been validated against high-quality administrative datasets [39]. Despite potential for bias, we are encouraged that colocation data approximated the proportional makeup of at-risk travel well and provided more fine-scale spatial data compared to mobile phone coverage, which can comprise a variety of malaria risk regions [8].

In summary, malaria importation to Zanzibar is driven by high-risk travel from a few mainland regions, influenced by seasonal and occupational mobility. Targeted interventions informed by both mobility data and identified risk groups may offer an effective strategy for reducing importation.

## Data Availability

The clinic datasets analyzed during this study are available in the UNC Dataverse repository, https://doi.org/10.15139/S3/F6FKRL. Colocation and movement distribution data are available from Meta Data For Good, but restrictions apply to the availability of these data and are not publicly available.

## LIST OF ABBREVIATIONS

### Abbreviation Definition

CHIRPS: Climate Hazards Group InfraRed Precipitation with Station data
CRF: Case Report Form
DFG: Meta Data For Good
GADM: Database of Global Administrative Boundaries
IRB: Institutional review board
IRR: Incidence rate ratio
MUHAS: Muhimbili University of Health and Allied Sciences
RDT: Rapid diagnostic test
UNC: University of North Carolina
ZAHREC: Zanzibar Health Research Ethics Committee
ZAMEP: Zanzibar Malaria Elimination Program
ZIM: Zanzibar Imported Malaria

## DECLARATIONS

### Ethics approval and consent to participate

Ethical approval was obtained from the institutional review boards (IRB) in Zanzibar (ZAHREC/01/EXT/July/2023/03), at MUHAS (MUHAS-REC-02-2024-1168), and at UNC (21-1966). Written informed consent, parental consent, and/or assent were provided by all participants.

### Consent for publication

Not applicable.

### Authors’ contributions

Conception and design: JTL, JJJ, VG, BEN, RR, AB, ME, DB; Data acquisition, analysis, and interpretation: JGM, KD, VG, MA, WH, BM, BBC, DM, AZ, MN, RK, SJS, JTL; Drafting manuscript: JGM, KDM, JTL, RR; Revision and final approval: All authors; Accountability: JGM, KD, BEN, JJJ, JTL.

## Acknowledgments

We thank the study participants who donated their time and the health facility staff who collected samples for the study. We thank the ZAMEP district malaria surveillance officers who were instrumental to the successful implementation of the study.

