## Supplemental Material for "Integrating mobility, travel survey, and malaria case data to understand drivers of malaria importation to Zanzibar, 2022-2023"

### Supplemental Methods

**Meta Colocation Data**
Meta calculates the colocation rate as the quotient of the measured co-observation and measured colocation rates. The co-observation rate is the probability that two individuals are simultaneously observed, while the colocation rate is the probability that two individuals are observed at the same time and in the same location. Meta does not release data on the counts behind the proportions, so there is no known sample size, other than the privacy floor of 10 people per district per day.

The data were first cleaned by removing district-district pairs with three or fewer weekly observations. Observations where either the measured co-observation or measured colocation rates were outliers (>1.5*IQR outside of IQR) for that district-district pair were then dropped.

### Table S1

**Table S1. Name, district, and number of cases from each of the selected Zanzibar health clinics.**

| Clinic ID | Clinic Name | District | Cases (n) |
|---|---|---|---|
| ZB_C001 | Jumbi Dispensary | Kati | 94 |
| ZB_C002 | Chukwani | Kati | 126 |
| ZB_C003 | Mafunzo | Mjini | 94 |
| ZB_C004 | Bumbwisudi | Magharibi A | 63 |
| ZB_C005 | Jangombe Mpendae | Mjini | 190 |
| ZB_C006 | Raha Leo | Mjini | 155 |
| ZB_C007 | Kianga PHCU | Magharibi A | 58 |
| ZB_C008 | Al Tabib Dispensary | Mjini | 44 |
| ZB_C009 | Utapoa Dispensary | Mjini | 102 |
| ZB_C010 | Matrekta | Magharibi B | 21 |
| ZB_C011 | Mwera | Kati | 71 |
| ZB_C012 | Kidongo Chekundu | Mjini | 46 |
| ZB_C013 | Makunduchi | Kusini | 83 |
| ZB_C014 | Kitogani | Kusini | 14 |
| ZB_C015 | Magogoni | Magharibi B | 70 |
| ZB_C016 | Kiembe Samaki | Magharibi B | 53 |
| ZB_C017 | Kizimkazi Mkunguni | Kusini | 92 |
| ZB_C018 | Minna Dispensary | Magharibi A | 74 |
| ZB_C019 | Kitope Dispensary | Kaskazini B | 30 |
| ZB_C020 | Kisauni | Magharibi B | 63 |
| ZB_C021 | Jendele | Kati | 16 |
| ZB_C022 | Koani Dispensary | Kati | 3 |
| ZB_C023 | Kidimni PHCU | Kati | 69 |
| ZB_C024 | Ndijani Mseweni | Kati | 28 |
| ZB_C025 | Kauthar Specialized Clinic | Kaskazini A | 43 |
| ZB_C026 | Mkokotoni PHCU | Kaskazini A | 24 |
| ZB_C027 | Pwani Mchangani PHCU | Kaskazini A | 22 |
| ZB_C028 | Dr. Metha Nungwi | Kaskazini A | 1 |
| ZB_C029 | Upenja | Kaskazini B | 35 |
| ZB_C030 | Altwaybib Dispensary | Kaskazini B | 18 |
| ZB_C031 | Magirisi | Magharibi B | 94 |
| ZB_C032 | Bwefum | Magharibi B | 23 |
| ZB_C033 | Mtofaani PHCU | Magharibi A | 64 |
| ZB_C034 | Muyuni | Kusini | 24 |
| ZB_C035 | Mwera Pongwe | Kati | 17 |
| ZB_C037 | Afaa Medical Clinic | Magharibi B | 17 |
| ZB_C038 | Miwani | Kati | 42 |
| ZB_C039 | Kizimkazi Dimbani | Kusini | 54 |
| ZB_C040 | Bububu Dispensary | Magharibi A | 17 |
| ZB_C041 | Kendwa PHCU | Kaskazini A | 12 |
| ZB_C042 | Machui | Kati | 12 |
| ZB_C043 | Mchangani | Kati | 6 |
| ZB_C044 | Al-Manna Dispensary | Kaskazini A | 15 |
| ZB_C045 | Ubago Jeshini | Kati | 14 |
| ZB_C046 | Charawe | Kati | 36 |
| ZB_C047 | Bwejuu | Kusini | 15 |
| ZB_C048 | Chwaka | Kati | 18 |
| ZB_C049 | Bumbwini Misufini | Kaskazini B | 9 |

| | | | |
|---|---|---|---|
| ZB_C050 | Nour Alaa Nour | Kaskazini A | 94 |
| ZB_C051 | Mnazi Mmoja | Mjini | 11 |
| ZB_C052 | Azhar | Magharibi A | 21 |
| ZB_C053 | Triple J Medical Services | Magharibi B | 58 |
| ZB_C054 | Kombeni | Magharibi B | 12 |
| ZB_C055 | Kundi Dispensary | Mjini | 32 |
| ZB_C056 | Shash Dispensary | Magharibi B | 67 |
| ZB_C057 | Icb Dispensary | Mjini | 19 |
| ZB_C058 | Habiba Dispensary | Magharibi A | 39 |
| ZB_C059 | Bandarini | Mjini | 33 |
| ZB_C060 | Mwembeladu Hospital | Mjini | 38 |
| ZB_C061 | Cheju | Kati | 102 |
| ZB_C062 | Gana | Kati | 6 |
| ZB_C063 | Kiboje | Kati | 6 |
| ZB_C064 | Marumbi | Kati | 5 |
| ZB_C065 | Ndijani Baniani | Kati | 7 |
| ZB_C066 | Jendele | Kati | 20 |
| ZB_C067 | Uzini | Kati | 35 |
| ZB_C068 | Pongwe | Kati | 6 |
| ZB_C069 | Umbuji | Kati | 22 |
| ZB_C070 | Ahsana Dispensary | Kaskazini A | 4 |
| ZB_C071 | Kivunge | Kaskazini A | 31 |
| ZB_C073 | Kijini PHCU | Kaskazini A | 12 |
| ZB_C074 | Nungwi PHCU+ | Kaskazini A | 24 |
| ZB_C075 | Al-Shifaa Kazole Dispensary | Kaskazini B | 33 |
| ZB_C076 | Fujoni | Kaskazini B | 15 |
| ZB_C078 | Kibuteni | Kusini | 4 |
| ZB_C079 | Michamvi | Kusini | 14 |
| ZB_C080 | Hassan Clinic | Mjini | 10 |
| ZB_C081 | J And M Dispensary | Mjini | 14 |
| ZB_C082 | Kidutani | Mjini | 92 |
| ZB_C083 | Ziwani Polisi | Mjini | 175 |
| ZB_C084 | New Asaakheri | Mjini | 58 |
| ZB_C085 | Shaurimoyo | Mjini | 97 |
| ZB_C087 | Al-Najid | Magharibi A | 6 |
| ZB_C088 | Kizimbani PHCU | Magharibi A | 26 |
| ZB_C089 | Mbuzini Clinic And Dispensary | Magharibi A | 51 |
| ZB_C092 | Znz Clinic | Magharibi A | 28 |
| ZB_C093 | Al-Hijri | Magharibi B | 2 |
| ZB_C094 | Sanasa | Magharibi B | 83 |
| ZB_C095 | Al-Majid Dispensary | Magharibi B | 1 |
| ZB_C096 | Care Clinic | Magharibi B | 90 |
| ZB_C097 | Marie Stop Hospital | Magharibi B | 14 |
| ZB_C098 | Mayda Health Charitable | Magharibi B | 10 |
| ZB_C099 | Sahal Hospital | Magharibi B | 55 |
| ZB_C100 | St. Camiliusn Dispensary | Magharibi B | 2 |

### Figure S1

**Figure S1. Region- (A) and district-level (B) malaria risk stratification of mainland Tanzania, according to the Tanzania National Malaria Strategic Plan.** Malaria risk strata were assigned based on five key indicators (parasite prevalence, fever test positivity rate, annual parasite incidence, confirmed malaria incidence, and malaria positivity in pregnant women). Districts were categorized into four risk levels (very low, low, moderate, high) using a conservative approach that classified each district by its highest recorded risk over the past three years. Districts and regions are colored according to assigned risk stratum. Taken from the Tanzania National Malaria Strategic Plan, 2021-2025.

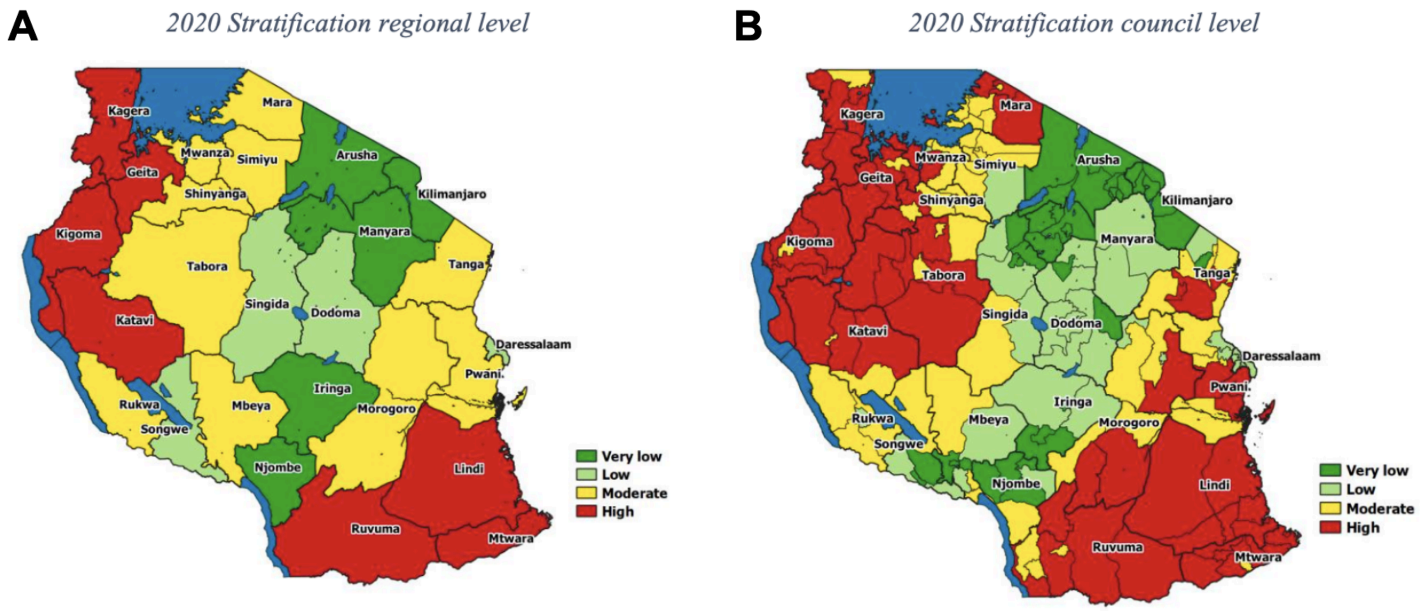

### Table S2

**Table S2. Select demographic characteristics of confirmed malaria cases on Zanzibar who did not report travel to the mainland within the past 28 days, 2022-2023.** This table presents self-reported sex, age, and occupation for clinic cases captured in Zanzibar (overall and stratified by self-reported travel within Zanzibar). Differences between travelers and non-travelers within each stratum were assessed using Fisher's exact test or chi-squared test, as appropriate. Bolded p-values are significant (*p*<0.05).

| | | Travel Within Zanzibar | | |
|---|---|---|---|---|
| | **Total (N=2582)** | **No (N=2109)** | **Yes (N=473)** | **P-value** |
| **Sex** | | | | |
| Female | 745 (28.9%) | 619 (29.4%) | 126 (26.6%) | 0.49 |
| Male | 1835 (71.1%) | 1488 (70.6%) | 347 (73.4%) | |
| **Age** | | | | |
| Under 5 | 97 (3.8%) | 84 (4.0%) | 13 (2.7%) | 0.54 |
| 5 to 14 | 269 (10.4%) | 231 (11.0%) | 38 (8.0%) | |
| 15 to 24 | 929 (36.0%) | 759 (36.0%) | 170 (35.9%) | |
| 25 to 39 | 847 (32.8%) | 675 (32.0%) | 172 (36.4%) | |
| 40 and older | 440 (17.0%) | 360 (17.1%) | 80 (16.9%) | |
| **Occupation*** | | | | |
| Trader/business | 522 (25.3%) | 450 (26.9%) | 72 (18.3%) | **0.01** |
| Farming | 213 (10.3%) | 172 (10.3%) | 41 (10.4%) | 0.94 |
| Housewife | 209 (10.1%) | 185 (11.1%) | 24 (6.1%) | **0.03** |
| Watchman/security | 205 (9.9%) | 146 (8.7%) | 59 (15.0%) | **<0.001** |
| Student | 171 (8.3%) | 129 (7.7%) | 42 (10.7%) | 0.10 |
| Fishing | 130 (6.3%) | 99 (5.9%) | 31 (7.9%) | 0.25 |
| Public servant/NGO | 94 (4.5%) | 70 (4.2%) | 24 (6.1%) | 0.19 |
| Tourism | 63 (3.0%) | 44 (2.6%) | 19 (4.8%) | **0.05** |
| Food service | 67 (3.2%) | 54 (3.2%) | 13 (3.3%) | 0.98 |
| Construction | 33 (1.6%) | 29 (1.7%) | 4 (1.0%) | 0.65 |
| Other | 359 (17.4%) | 295 (17.6%) | 64 (16.3%) | 0.96 |
| Child | 488 | 412 | 76 | |
| Missing | 28 | 24 | 4 | |

*Percentages are based on totals minus children and those missing occupation.

### Figure S2

**Figure S2. Distribution of nights spent at destination among confirmed Zanzibari malaria cases who reported recent travel to the mainland and a surveyed sample of outbound ferry travelers at the Dar es Salaam ferry terminal, 2022-2023.** The x-axis represents categorical time intervals of nights spent at destination, as presented on the survey provided to clinic cases and ferry travelers, while the y-axis represents the number of travelers in each category. 171 clinic case travelers were missing data for the number of nights spent away, and are excluded from the plot.

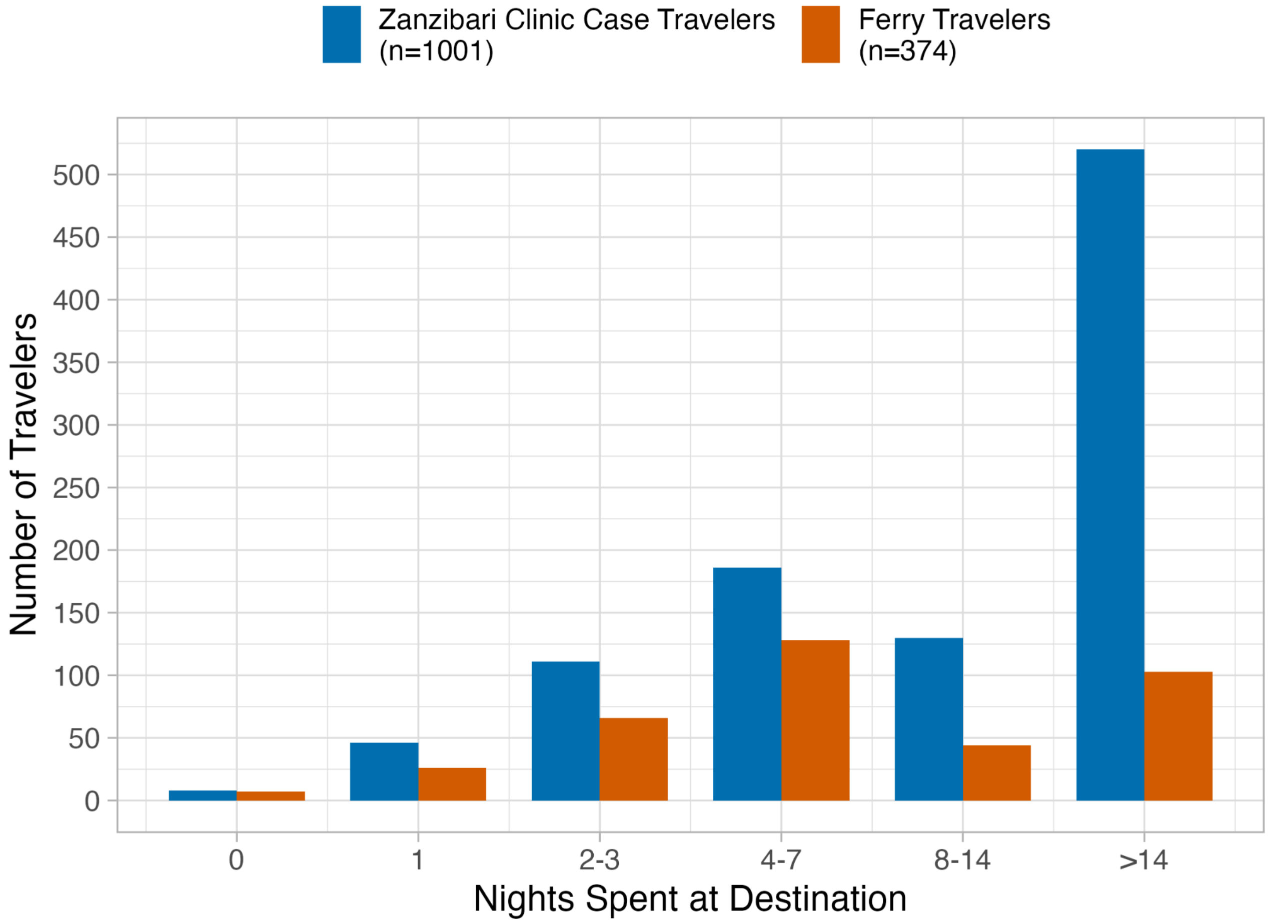

### Table S3

**Table S3. Select demographic characteristics of confirmed malaria cases on Zanzibar who reported mainland Tanzania to be their primary residence (n=600), 2022-2023.** This table presents self-reported gender, age, occupation, and recent travel to the mainland for mainland-residing clinic cases captured in Zanzibar (overall, stratified by self-reported months spent on Zanzibar, and stratified by recent travel to the mainland).

| | | | No Recent Travel | |
|---|---|---|---|---|
| | Total (N=600) | Recent Travel (N=333) | ≤6 Months on ZB (N=116) | >6 Months on ZB (N=151) |
| **Sex** | | | | |
| Female | 252 (42.0%) | 146 (43.8%) | 46 (39.7%) | 60 (39.7%) |
| Male | 348 (58.0%) | 187 (56.2%) | 70 (60.3%) | 91 (60.3%) |
| **Age** | | | | |
| Under 5 | 56 (9.3%) | 43 (12.9%) | 9 (7.8%) | 4 (2.6%) |
| 5 to 14 | 78 (13.0%) | 39 (11.7%) | 6 (5.2%) | 33 (21.9%) |
| 15 to 24 | 256 (42.7%) | 134 (40.2%) | 59 (50.9%) | 63 (41.7%) |
| 25 to 39 | 163 (27.2%) | 93 (27.9%) | 37 (31.9%) | 33 (21.9%) |
| 40 and older | 47 (7.8%) | 24 (7.2%) | 5 (4.3%) | 18 (11.9%) |
| **Occupation*** | | | | |
| Trader/business | 108 (24.0%) | 51 (21.2%) | 19 (19.0%) | 38 (34.9%) |
| Farming | 100 (22.2%) | 68 (28.2%) | 24 (24.0%) | 8 (7.3%) |
| Housewife | 52 (11.6%) | 29 (12.0%) | 9 (9.0%) | 14 (12.8%) |
| Watchman/security | 24 (5.3%) | 9 (3.7%) | 10 (10.0%) | 5 (4.6%) |
| Student | 24 (5.3%) | 10 (4.1%) | 4 (4.0%) | 10 (9.2%) |
| Fishing | 22 (4.9%) | 12 (5.0%) | 3 (3.0%) | 7 (6.4%) |
| Tourism | 22 (4.9%) | 11 (4.6%) | 7 (7.0%) | 4 (3.7%) |
| Public servant/NGO | 15 (3.3%) | 6 (2.5%) | 5 (5.0%) | 4 (3.7%) |
| Food service | 15 (3.3%) | 10 (4.1%) | 2 (2.0%) | 3 (2.8%) |
| Construction | 14 (3.1%) | 8 (3.3%) | 5 (5.0%) | 1 (0.9%) |
| Other | 54 (12.0%) | 27 (11.2%) | 12 (12.0%) | 15 (13.8%) |
| Child | 141 | 85 | 15 | 41 |
| Missing | 9 | 7 | 1 | 1 |

*Percentages are based on totals minus children and those missing occupation.

### Figure S3

**Figure S3. Counts (A) and proportions (B) of Zanzibari confirmed malaria cases who reported recent travel (n=1,172), 2022-2023.** Onboarding of Zanzibar health clinics was not completed until October 2022. Resurgence of locally transmitted malaria in Zanzibar in 2023 drove down the proportion of cases which traveled.

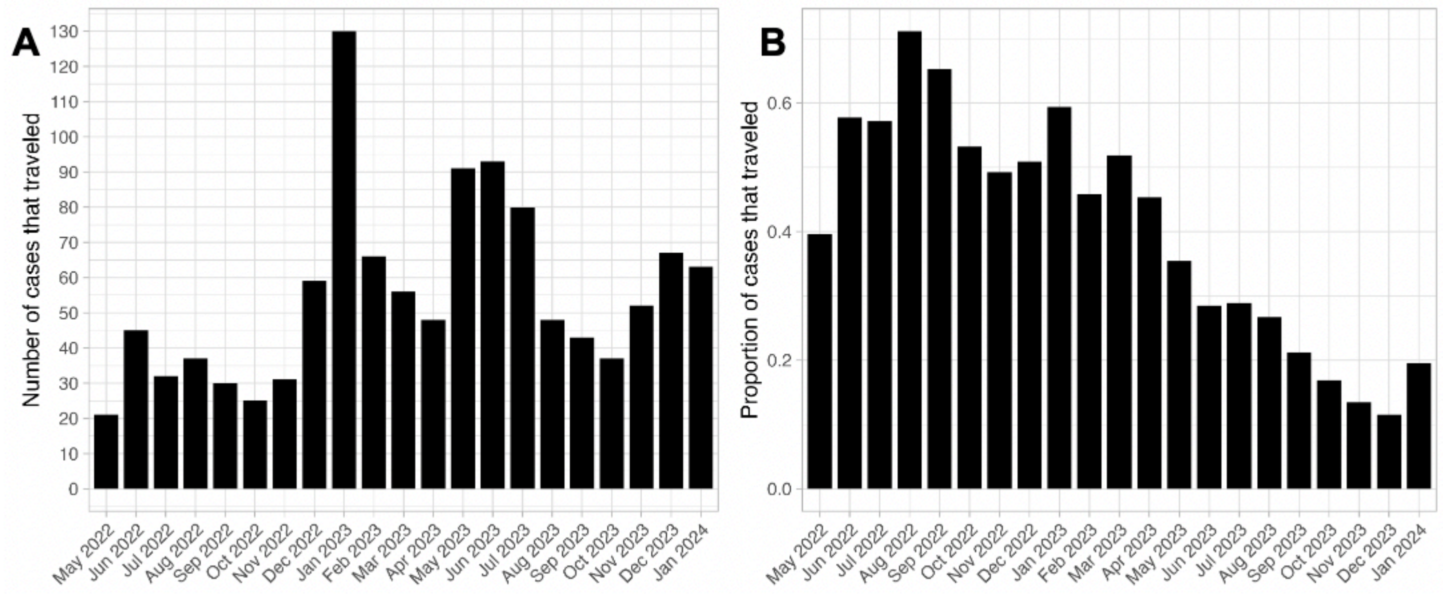

### Figure S4

**Figure S4. Colocation rates of mainland districts with each district in Unguja.** The top 7 outliers are labelled and colored by malaria risk stratification (red: high, yellow: moderate, green: very low) (Figure S1). Colocation is not weighted by the district's population. Pangani is the highest mainland district for colocation for all but Kati and Kusini.

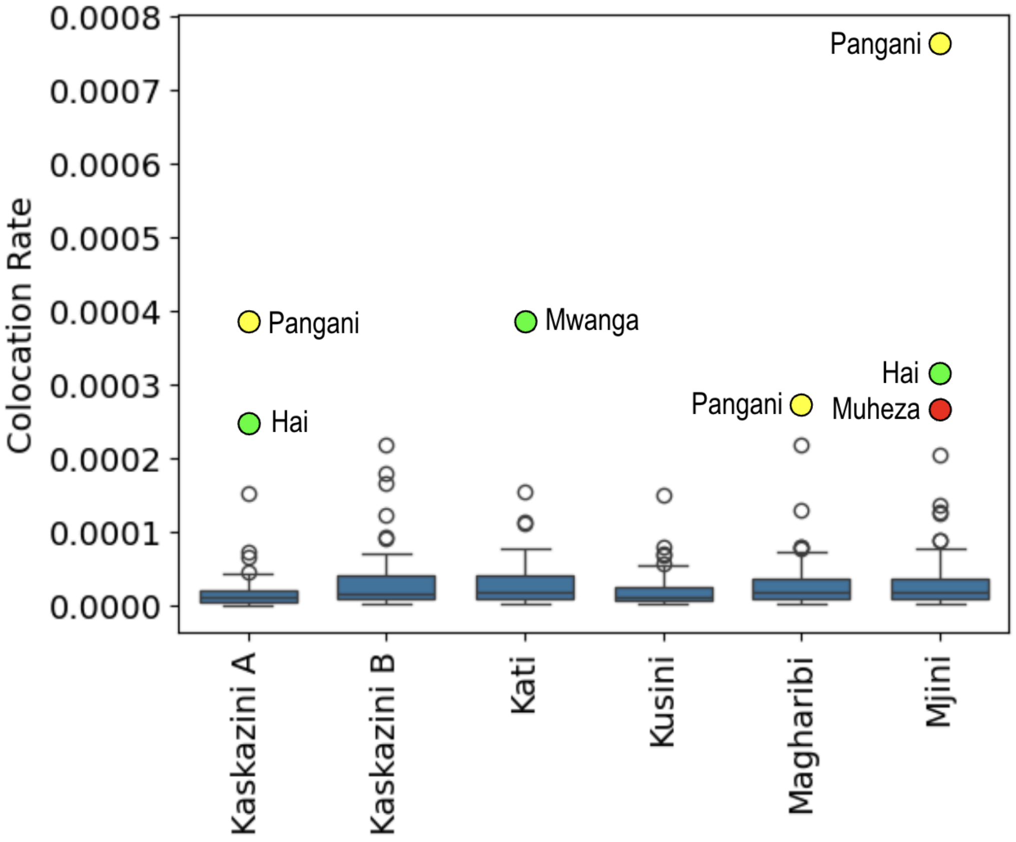

### Figure S5

**Figure S5. Colocation between Unguja and mainland regions, averaged over the wet (March-May, October-December) and dry months, 2022-2023.** We did not find that colocation differed by season when comparing colocation across weeks grouped by season using Mann Whitney-U testing.

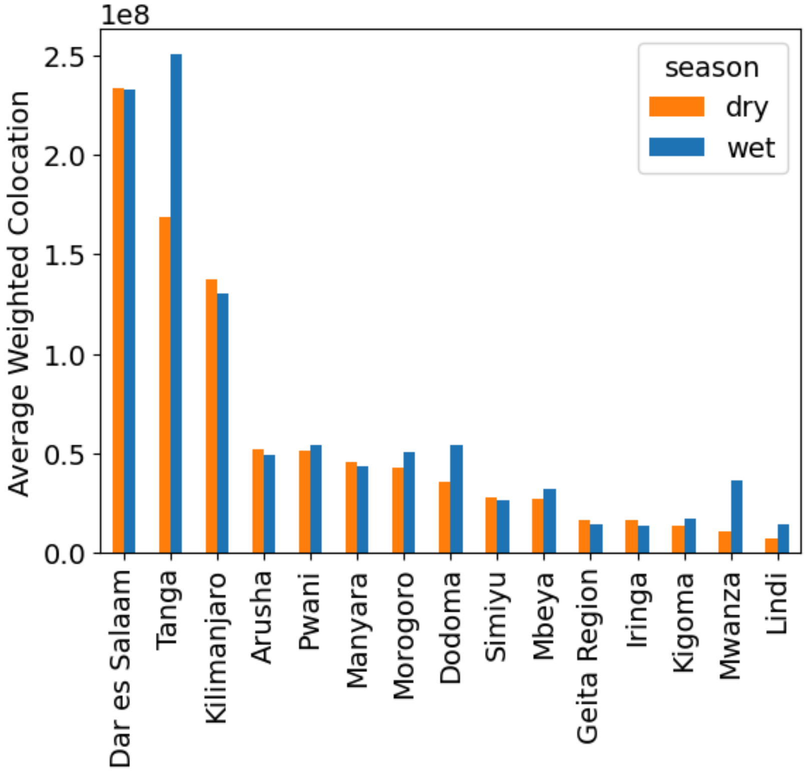

Panels A and B below depict weekly colocation data over time for two Unguja districts with the Tanga region on the mainland, stratified by weeks falling in dry vs wet season months. We did not detect differences in colocation by season using Mann Whitney-U testing.

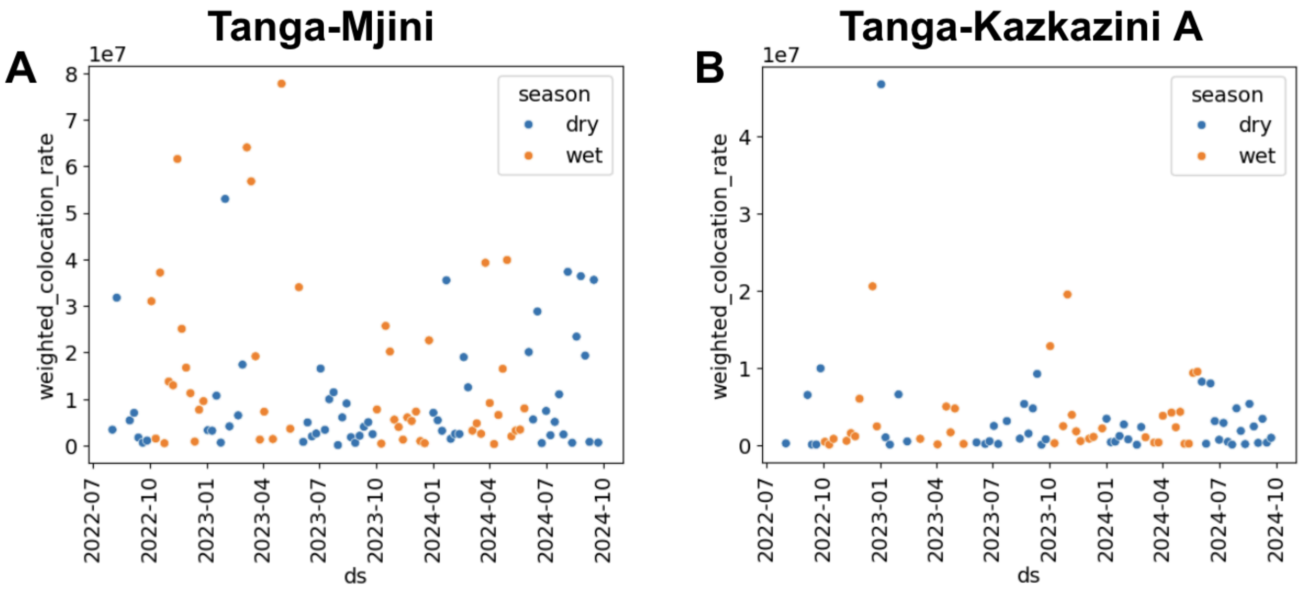

### Figure S6

**Figure S6. Counts (A and B) and proportions (C and D) of travel person-days for mainlanders vs Zanzibaris traveling to Zanzibar based on ferry passenger surveys.** (B)/(D) is the same figure as (A)/(C) but Dar es Salaam is omitted and the y-axis is rescaled.

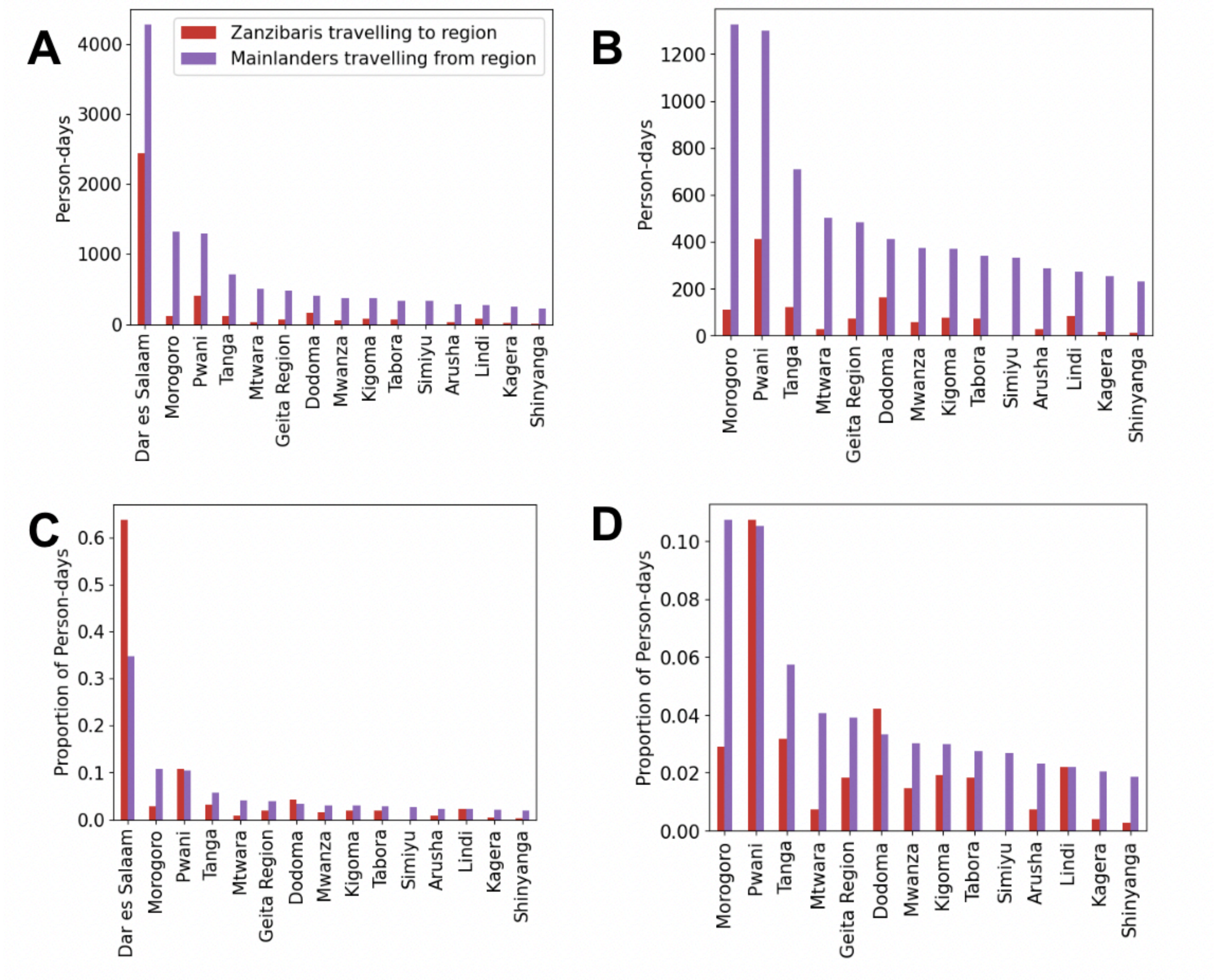

### Figure S7

**Figure S7. Proportion of contribution to colocation and clinic case data for each mainland region, among Zanzibar regions (Mjini Magharibi, Kaskazini Unguja, and Kusini Unguja), 2022-2023.** Note the difference in y-axes for each region. Approximately 30% of the Zanzibar locations for travelers in the ferry screening data were missing, so those data were omitted.

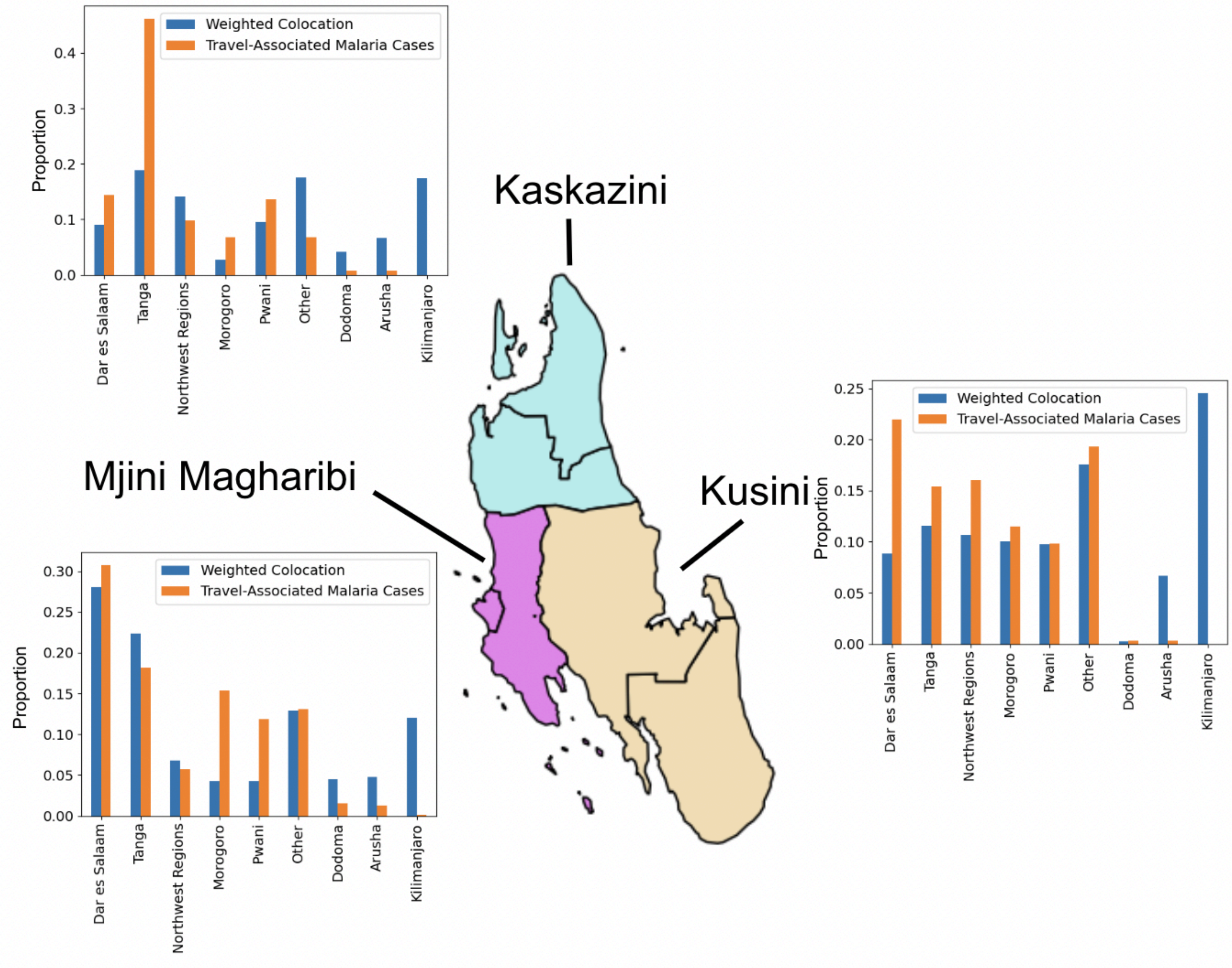

### Figure S8

**Figure S8. Comparison of ferry survey data and travel-associated malaria case data, stratified by mainlander/Zanzibari self-identification.** "Mainlander" clinic cases include individuals who presented to a Zanzibar health clinic with malaria, who indicated that their primary residence was on mainland Tanzania. Destinations for mainland clinic cases are either a self-reported recent travel destination, or their home district on the mainland (if no recent travel was reported and a mainland home district was provided).

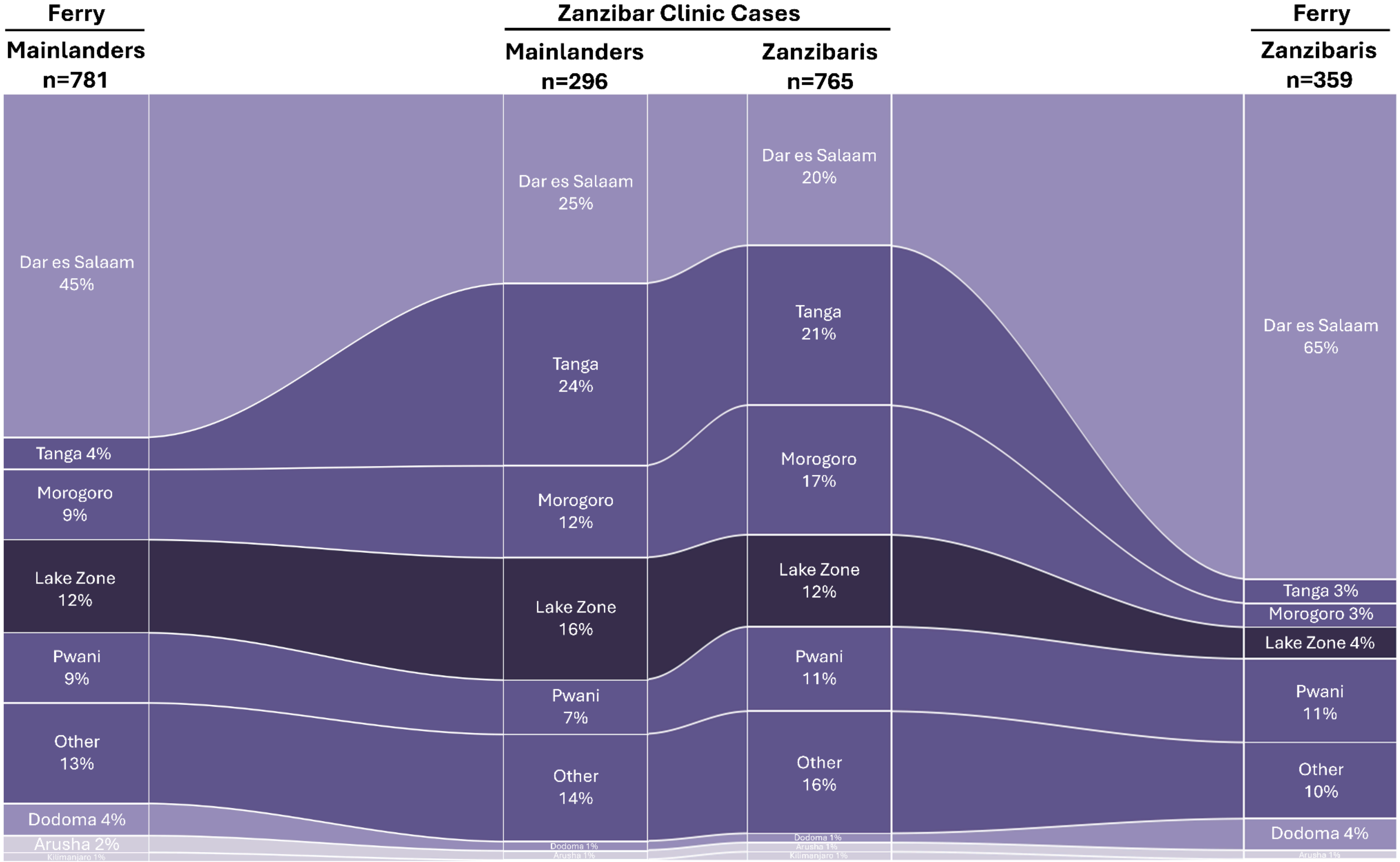
